# Oxygen and mortality in COVID-19 pneumonia: a comparative analysis of supplemental oxygen policies and health outcomes across 26 countries

**DOI:** 10.1101/2020.07.03.20145763

**Authors:** Daniel K Goyal, Harry Donnelly, Albrecht Kussner, James Neil, Sohail Bhatti, Fatma Mansab

## Abstract

**Introduction:** Hypoxia is the main cause of morbidity and mortality in COVID-19. During the COVID-19 pandemic some countries have reduced access to supplemental oxygen (e.g. oxygen rationing), whereas other nations have maintained and even improved access to supplemental oxygen. We examined whether such variation in the access to supplemental oxygen had any bearing on mortality in COVID-19.

**Methods:** Three independent investigators searched for, identified and extracted the nationally recommended target oxygen levels for the commencement of oxygen in COVID-19 pneumonia from the 29 worst affected countries. Mortality estimates were calculated from three independent sources. We then applied linear regression analysis to examine for potential association between national targets for the commencement of oxygen and case fatality rates.

**Results:** Of the 26 nations included, 15 had employed conservative oxygen strategies to manage COVID-19 pneumonia. Of them, Belgium, France, USA, Canada, China, Germany, Mexico, Spain, Sweden and the UK guidelines advised commencing oxygen when oxygen saturations (SpO2) fell to 91% or less. Target SpO2 ranged from 92% to 95% in the other 16 nations. Linear regression analysis demonstrated a strong inverse correlation between the national target for the commencement of oxygen and national case fatality rates (Spearman’s Rho = −0.622, p < 0.001).

**Conclusion:** Our study highlights the disparity in oxygen provision for COVID-19 patients between the nations analysed, and indicates such disparity in access to supplemental oxygen may represent a modifiable factor associated with mortality during the pandemic.

**Key Messages:** *What is already known?:*

- There were no prospective clinical trials we could identify relating to COVID-19 and supplemental oxygen, nor any published studies examining access to supplemental oxygen and mortality in COVID-19.
- There are a number of studies identifying an association with low oxygen saturations at presentation and mortality in COVID-19 pneumonia.
- There is good quality evidence that a delay in the correction of hypoxia in pneumonia increases mortality.

*What are the new findings?:*

- This study highlights the different thresholds for commencing supplemental oxygen in patients with COVID-19 across 26 nations.
- Those countries that provide better access to supplemental oxygen have a statistically significant lower mortality rate.
- Our results support the consensus view that improving access to supplemental oxygen in COVID-19 pneumonia is likely to reduce mortality.

## Introduction

SARS-CoV2 causes COVID-19 (Coronavirus Disease 2019). As of May 2020, the total reported cases of COVID-19 was over 5 million, with 350,000 deaths over five months[1]. More than half these deaths have occurred in the last month. Whilst there has been a slight reduction in the rate of growth for new infections globally, this is most likely due to strict infection control policies (e.g. case-isolation, social distancing and ‘lockdown’)[2]. With the seroprevalence of SARS-CoV2 being reported as between <1% to 22%[3], it is most likely the majority of infections are yet to come, and the rate of infections will once again increase as infection control measures are balanced with economic pressures.

The true COVID-19 mortality rate is difficult to ascertain during the outbreak. Background infections, asymptomatic infections, testing criteria, reporting of fatalities and the time-lag between new cases and outcome are all potential confounders[4]. This makes measuring the effects of national interventions difficult. It is though, reasonable to expect a nation’s COVID-19 mortality rate will depend on access to healthcare, and likely will also depend on the type of healthcare offered. The need for effective healthcare can be reasonably inferred from the marked disparity between mortality rates during a surge of cases versus mortality post-surge[5].

Oxygen is a cornerstone of treatment for patients with COVID-19 pneumonia. Indeed, the major mechanism for injury and death in COVID-19 relates to hypoxia[6]. Despite the critical nature of oxygen therapy in COVID-19 pneumonia there remains marked variation between national guidelines for when to offer supplemental oxygen. Many nations seem to have implemented conservative oxygen strategies during the pandemic, effectively limiting the access of patients to supplemental oxygen. Others seem to have actively increased their capacity to offer supplemental oxygen for patients with COVID-19 pneumonia. Here, we examine the national guidelines from 29 nations in an effort to understand the potential impact the varying thresholds for commencing supplemental oxygen have on COVID-19 outcomes.

## Methods

We followed the advice for global reporting on health estimates as per the GATHER statement[7]. All countries with more than 20,000 cases as of 18/05/20, were assessed. Three investigators independently identified the specific national recommendations for the target oxygen saturations (SpO2) to commence oxygen in patients with COVID-19. Two investigators were blinded as to the reason for the study. Each nation’s ministry of health, national guideline bodies, respiratory medicine bodies and national health service were searched for relevant COVID-19 clinical guidelines. The European Society of Respiratory Medicine was a useful resource with direct links to a number of COVID-19 specific clinical guidelines from across the world. Literature databases were also used as a means of identifying links to national guidelines. If guidelines were not available in one of the languages spoken by the investigators, on-line translation services were utilised, specifically for guidelines on ‘supplemental oxygen’ or ‘oxygen therapy’ - the entire guideline was not translated. Note, only guidelines applicable to the majority of the population were extracted, and guidelines for patients with underlying conditions such as chronic obstructive airways disease were not recorded.

If guidelines were unclear, instruction was to disregard the country from further analysis. Where there were more than one recommendation the investigator was to make a determination as to the most likely guideline to be followed (figure 1). Where there was divergence between the three investigators, the consensus value was used. Results were tabulated and compared.

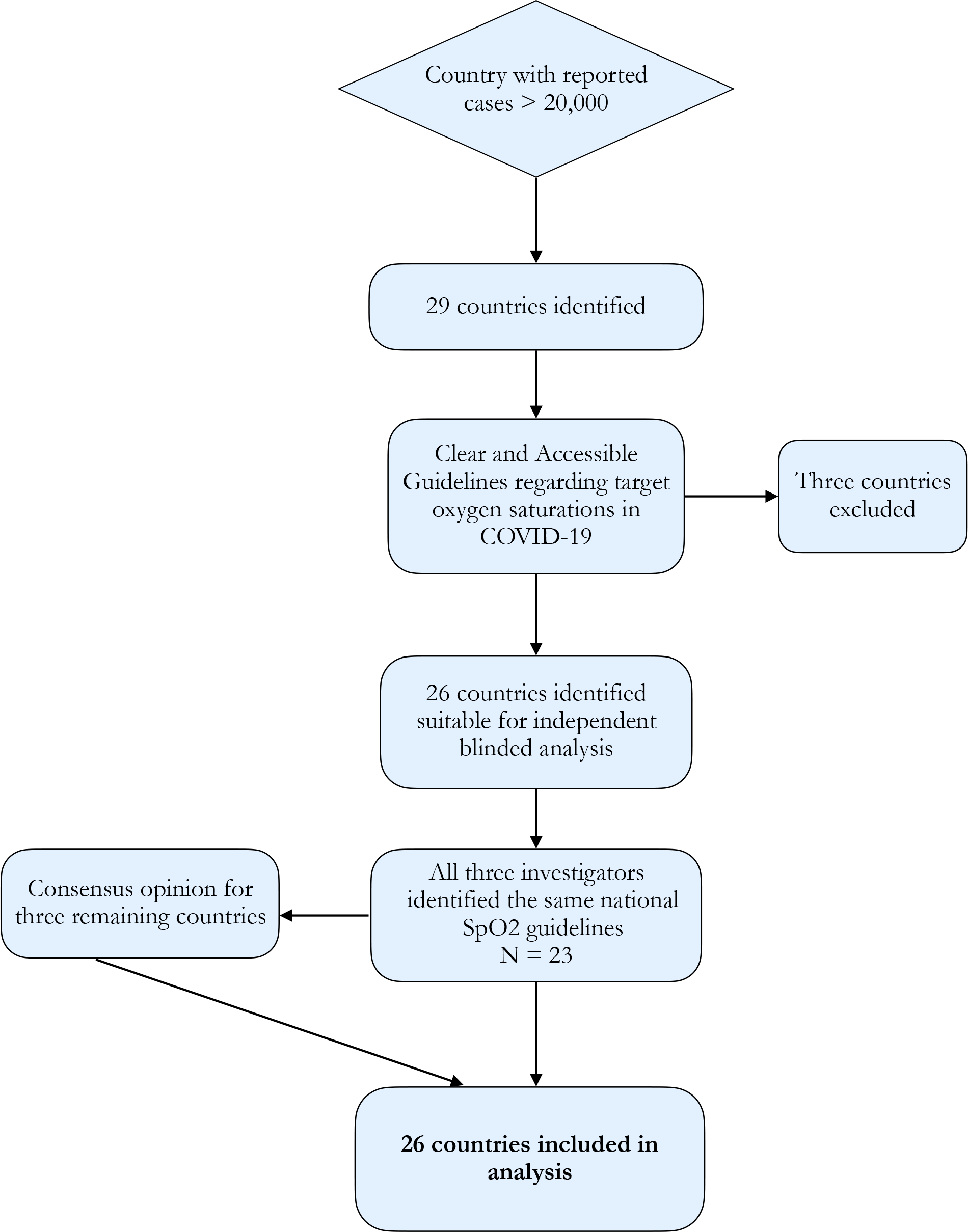

### Case Fatality Rate and Infection Fatality Rate

Case Fatality Rate (CFR) is the percentage ratio of deaths to total cases. It is a crude figure privy to a number of potential confounders. For most nations it is likely to be numerically incorrect[4]. CFR is though, likely to maintain a relationship to actual infection mortality rate (IFR)[3,4], and as such was used in this study. CFR was calculated and cross-referenced from three different sources - The WHO, John Hopkins University and Worldometer. There was no significant difference between the calculated CFR across the three sources.

### Patient and Public Involvement

It was not possible to include patients or public in the present study.

### Statistical Analysis

Linear regression was performed to identify a potential trend between CFR and Target SpO2, and presented using scatter-plots. Due to the sample size (n=26), it was not clear whether the assumptions of normality and linearity were met, so the statistical significance of the possible relationship between CFR and target SpO2 was established using the non-parametric Spearman’s-Rho test. We have also explored the effect of potentially confounding variables using the non-parametric Spearman’s-rho test rather than using a parametric Mancova test to adjust for confounding, given the small sample size and uncertainty regarding linearity and normality.

## Results

In total there were 29 countries with total case numbers over 20,000 on 18th May 2020. Of those, 26 countries had accessible clinical guidelines referring to target oxygen levels for the commencement of supplemental oxygen in COVID-19. UAE (United Arab Emirates) was excluded from further analysis as the national guidelines advised (at page 9) admitting all patients with COVID-19 to hospital, and commencing oxygen when ‘needed’ [8] - the country’s low CFR is noted. The Netherlands and Belarus were also excluded due to all three investigators failing to find clear national guidelines regarding oxygen targets.

Of the remaining 26 countries there was concordance between all three investigators identifying the same national target oxygen levels in 23 countries. Of the remaining 3 countries (UK, Pakistan and Qatar), determination of national target SpO2 in COVID-19 was made by consensus. For links to national guidelines please see Supplementary File.

Of the 26 nations analysed, six recommended commencing oxygen if SpO2 fell to below 95% (Singapore, Peru, Switzerland, Ireland, Qatar and Pakistan), five made recommendation for Below 94% (Saudi Arabia, Chile, Brazil, India and Russia), five for below 93% (Portugal, Iran, Turkey, Bangladesh and Italy), six for below 92% (Canada, Belgium, France, UK, USA and China) and four for below 91% (Germany, Mexico, Spain and Sweden). CFR ranged from 0·06% (Qatar) to 16·4% (Belgium). There was a strong inverse correlation between recommended target oxygen saturations and national case fatality rates (rs = −0·622, p < 0·001). A scatter graph with linear best-fit line is at Figure 2.

**Figure 2.**
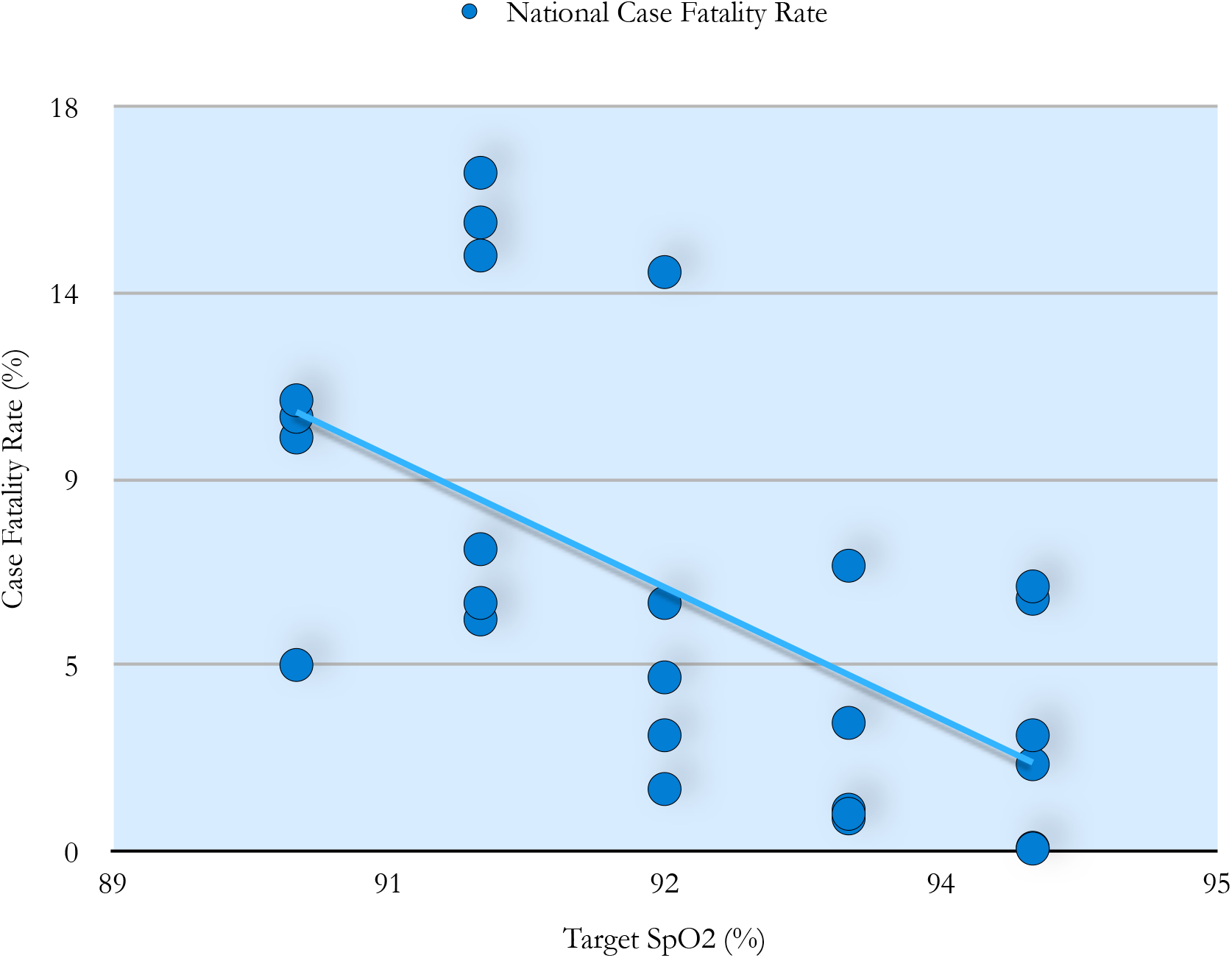
Scatter graph of national target oxygen saturations versus national case fatality rate with best fit linear line (n=26). SpO2 - oxygen saturations.

### Confounders

National guidelines for target saturations were relatively clear for most countries. Together with the high rate of consensus amongst investigators it seems unlikely that investigator bias was a significant factor. The main confounders are more likely to stem from the many variables associated with CFR.

We found no correlation between CFR and cases/million inhabitants (Figure 3B), or tests/ thousand inhabitants (Figure 3C), or overall positivity rate (Figure 3C), suggesting testing strategy between the countries examined did not have a significant visible relationship with our mortality measure, CFR (Table 1). We could not examine the potential impact of national-level reporting bias on the CFR from the data available.

**Table 1.** National CFR, target SpO2 and potential testing confounders in 26 countries. CFR - Case Fatality Rate; SpO2 - Oxygen Saturations. Overall Positivity Ratio-percentage ratio of positive to total cases; Case Burden - total number of positive cases per million inhabitants. Data extracted from WHO, Worldometer and John Hopkins University.

|  | SpO2 | Tests per<br>Thousand | Positivity<br>Ratio | Case Burden | CFR |
| --- | --- | --- | --- | --- | --- |
|  | (%) |  | (%) | (cases/<br>million) | (%) |
| **Qatar** | 94 | 82.3 | 25.9 | 20311 | 0.06 |
| **Singapore** | 94 | 57.2 | 10.5 | 6036 | 0.08 |
| **Pakistan** | 94 | 2.6 | 12.9 | 329 | 2.1 |
| **Peru** | 94 | 33.2 | 15.8 | 5163 | 2.8 |
| **Switzerland** | 94 | 46.3 | 7.7 | 3569 | 6.1 |
| **Ireland** | 94 | 70.6 | 7.7 | 5080 | 6.4 |
| **Saudi Arabia** | 93 | 24.6 | 10.6 | 2506 | 0.79 |
| **Russia** | 93 | 76.4 | 3.8 | 2843 | 0.9 |
| **Chile** | 93 | 32.1 | 17.5 | 5505 | 1 |
| **India** | 93 | 2.9 | 5.2 | 144 | 3.1 |
| **Brasil** | 93 | 4.4 | 56.9 | 2492 | 6.9 |
| **Bangladesh** | 92 | 2.0 | 15.4 | 301 | 1.5 |
| **Turkey** | 92 | 24.9 | 7.9 | 1955 | 2.8 |
| **Portugal** | 92 | 83.0 | 4 | 3206 | 4.2 |
| **Iran** | 92 | 11.6 | 16.1 | 1841 | 6 |
| **Italy** | 92 | 65.5 | 6 | 3857 | 14 |
| **China** | 91 | N/A | N/A | N/A | 5.6 |
| **USA** | 91 | 56.2 | 10.2 | 5620 | 6 |
| **Canada** | 91 | 45.7 | 5.4 | 2431 | 7.3 |
| **UK** | 91 | 68.0 | 6.4 | 4072 | 14.4 |
| **France** | 91 | 21.2 | 13.7 | 2899 | 15.2 |
| **Belgium** | 91 | 76.3 | 6.7 | 5051 | 16.4 |
| **Germany** | 90 | 47.1 | 4.6 | 2194 | 4.5 |
| **Spain** | 90 | 86.9 | 7 | 6133 | 10 |
| **Mexico** | 90 | 2.2 | 34.4 | 725 | 10.5 |
| **Sweden** | 90 | 27.3 | 15.8 | 3746 | 10.9 |
| **Correlation with CFR**<br>**(Spearman's Ro)** | **-0.622** | 0.0658 | -0.13 | 0.018 |  |
| **2-tailed (p-value)** | **<0.001** | 0.75 | 0.54 | 0.93 |  |

**Figure 3.**
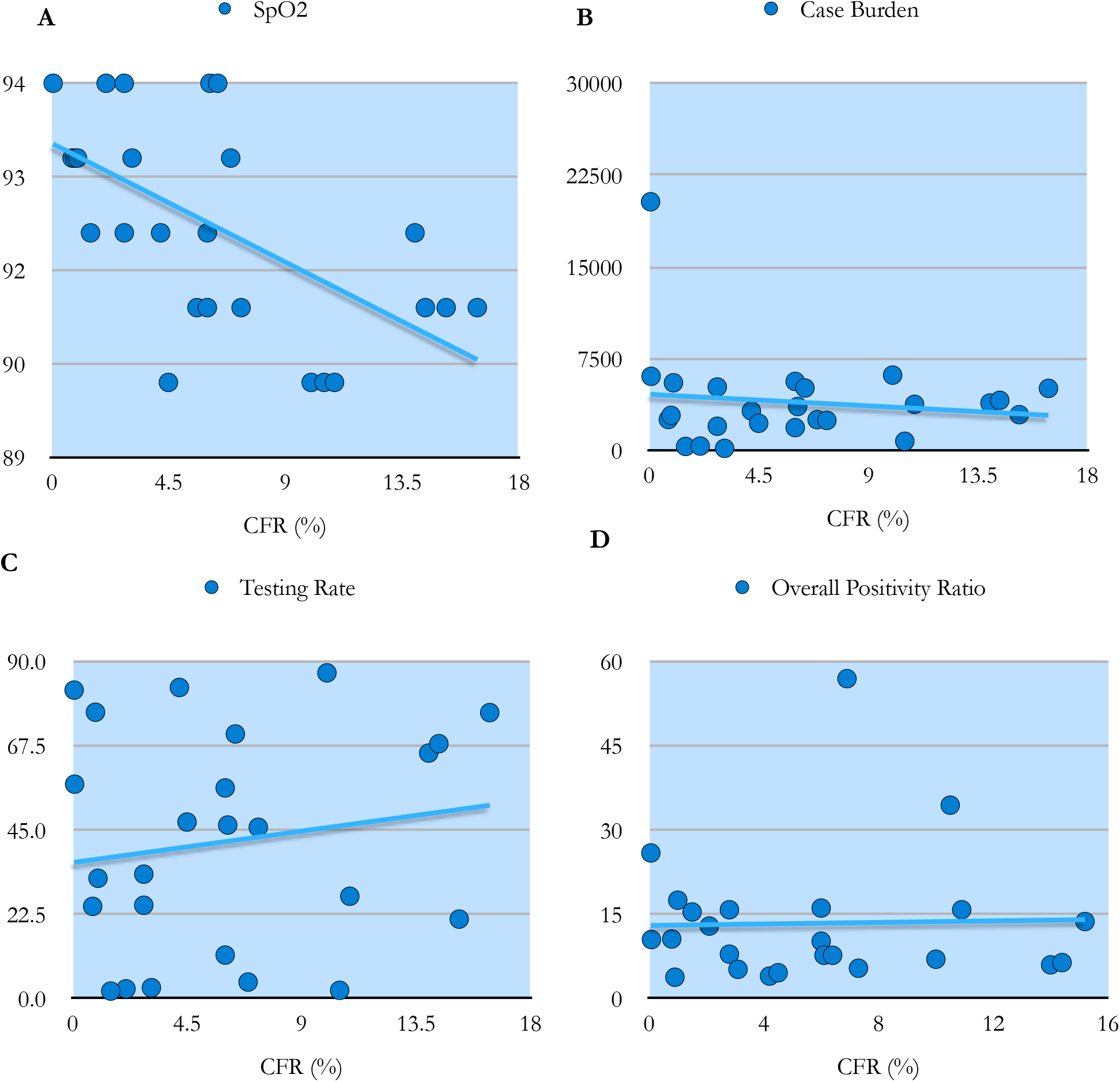
Scatter graphs with best-fit line for primary measure and confounders versus CFR (N= 26). CFR is along the horizontal axis in each and measured in percentage. A) Comparison of national CFR versus nationally recommended target SpO2 (%) in COVID-19. B) CFR versus burden of cases measured by total number of cases per million. C) CFR versus Testing Rate measured by total number of tests undertaken per thousand inhabitants (tests/ thousand). D) CFR versus the overall positivity ratio (%), measured as the percentage ratio of positive tests to total tests. CFR - Case Fatality Rate; SpO2 - oxygen saturations. CFR - Case Fatality Rate; SpO2 - Oxygen Saturations.

## Discussion

National guidelines for when to commence supplemental oxygen in patients with COVID-19 varied significantly between the countries examined. Combined, the target SpO2 for the *commencement* of oxygen and target SpO2 for ongoing treatment varied from 90% to 98%. Countries that implemented conservative oxygen strategies in response to the pandemic - effectively limiting access to supplemental oxygen - had statistically significant higher case fatality rates. There are a number of potential reasons for the variation in oxygen policies between countries and the association with mortality.

### Causative effect

It has been established that a delay in identifying and correcting hypoxia in pneumonia leads to increased disease severity, increased rate of mechanical ventilation and increased mortality[9,10]. Whilst there are no controlled studies in COVID-19 specifically examining duration spent hypoxic and subsequent disease burden and mortality, Sun et al., has reported a reduction in the need for mechanical ventilation where hypoxia was detected and corrected early in patients with COVID-19[11].

In relation to conservative oxygen strategies generally, there are four key mortality studies. The IOTA meta-analysis published in 2018 examined conservative versus liberal oxygen strategies across a range of studies. Whilst none of the studies analysed in the IOTA meta-analysis related to pneumonia, and the majority of studies examined oxygen as a treatment *not* as a means to correct hypoxia, the authors suggest optimal target SpO2 for all acute medical patients might be 94-96%[12].

Since the IOTA study, there have been three clinical studies, two of which were randomised controlled trials (RCT), examining the mortality effect of conservative oxygen. The ICU-ROX trial suggests there may be no mortality effect at the higher target levels of SpO2 (96-97% versus 95-96%) in mechanically ventilated patients from any cause (n=1000)[13]. Another, retrospective analysis, published in March 2020 examined over 35,000 intensive care patients and found the optimum SpO2 target of 94-98%. The authors note that patients who were in the optimal range for only 40% of the time had nearly twice the mortality of those who spent 80% of the time within the optimal target[14].

The most recent RCT, and the most well-controlled study of true conservative oxygen strategies to date (and the most relevant to COVID-19), examined 204 patients with acute respiratory distress syndrome (ARDS). Patients were randomised to either a conservative arm (actual SpO2 of 92-93%) versus a liberal arm (SpO2 of 95-97%), and then followed up for 90 days. The study was halted early due to excessive deaths in the conservative oxygen group, with a 27% increase in intensive care deaths and a 50% increase in 90 day mortality[15].

Based on our current understanding of the affects of hypoxia on inflammation[16] and coagulation[17], there is good scientific basis for the increased mortality associated with sub-optimal oxygen strategies and/or a delay in correcting hypoxia. There are direct effects of hypoxia leading to increased mortality, such as cardiac arrhythmias and ischaemic related pathologies (as identified in the aforementioned ARDS study[15]). It is also quite plausible, indeed quite likely, given that hypoxia is pro-inflammatory, the delay in correcting hypoxia leads to more severe disease. This of course raises the possibility that rationing, or a conservative oxygen approach, or a failure to provide access to supplemental oxygen in COVID-19 pneumonia, actually increases healthcare burden and resource consumption.

Whilst sometimes arguably unavoidable, the decision to limit access to supplemental oxygen should be undertaken mindful of the likely mortality impact, and of the possibility of perpetuating a healthcare crisis. Implementing oxygen early is likely to prevent disease progression, as suggested by Sun et al. [11], and as is consistent with established practice relating to pneumonia generally [9,10].

### Resource limitations

For some nations there was a resource limitation issue, or at least a fear of resource limitation, with secondary implementation of conservative oxygen strategies. For example, the UK directive to ration oxygen supply in April 2020 reduced the normal national target for the commencement of oxygen from SpO2 of 94% to a new value of 91%. The reason for rationing was related to the surge of infections and subsequent concern over the supply of oxygen[18]. If such practises are common in other nations, the relationship between national guidelines’ SpO2 and national CFR identified here may be a representation of the demands on healthcare during a surge of COVID-19 cases.

There are a number of reasons why mortality increases during a surge of infections. Patients are less likely to attend hospital or seek medical care, either for fear of contracting COVID-19 or over-burdening their health service[19]. Triage systems during a surge can be set with high thresholds for onward referrals[20]. Another mortality factor is a potential lack of resources both staff and consumables. The overall delay to treatment that ensues prevents early correction of hypoxia, implementation of VTE (venous thromboembolism) prophylaxis, readjustment of medications (e.g. nephrotoxics) and the detection of secondary bacterial infection, and thus a likely increased mortality[21]

So then, the association between target SpO2 and CFR identified here may be more related to target SpO2 being an indicator of an overwhelmed healthcare service. If this is the case, a lower than usual target SpO2 may still contribute to the higher mortality experienced in nations that suffered an overwhelming surge. In a UK cohort, during the initial surge of infections, and during oxygen rationing, approximately 62% of patients presented hypoxic, and of those that died 75% (60/80) of patients presented hypoxic (defined broadly in this study as SpO2 <95%)[22]. In a cohort from Mexico, 96% of the patients who died presented with SpO2 under 88%[23].

Part of the increased mortality seen during a surge of infections may be related to the secondary conservative oxygen policies and delay in correction of hypoxia.

### National Approach

The issuing of national guidelines recommending lower target oxygen saturations than would be typical for viral pneumonias[24], may relate more to the overall approach of a national response to COVID-19, and as such, it is this ‘national approach’ that relates to mortality rate.

All three investigators noted the quite different approaches between nations, as set out in their national guidelines. Some followed a ‘stay home’ approach, whereas others defaulted to clinical assessment of patients either with COVID-19 or with any risk factor associated with it. For example, Singapore guidelines default to clinical assessment[25], whereas a country with a similar prevalence burden, the UK, has much higher thresholds for referral onward for assessment[26] (Table 2).

**Table 2.** Comparison of the Criteria for Assessment in suspected or confirmed COVID-19 between Singapore and the UK. Information is based on the clinical guidelines from each nation and WHO (see Supplementary File). SpO2 - oxygen saturations. CFR - case fatality rate.

|  | Singapore | UK |
| --- | --- | --- |
| **Criteria for Clinical Assessment** |  |  |
| SpO2 (%) | < 95 | < 92 |
| Age (yrs) | > 65 | Irrelevant |
| Co-morbidity | Any | Severe |
| Duration of illness (days) | > 3 | Irrelevant |
| **Epidemiology** |  |  |
| Cases/million inhabitants | 6,063 | 4,076 |
| Physicians/10,000 head of capita | 24 | 28 |
| CFR (%) | 0.08 | 13.4 |

In this situation, where the national guideline target SpO2 is part of an overall strategy of avoiding admissions, then whilst it does remain likely conservative oxygen approaches do contribute to higher mortality, there may also be contribution of other policies. In the UK versus Singapore example, a target SpO2 of less than 92% is likely to be harmful, but equally, failing to account for age of the patient or duration of fever may also be harmful. As such, the relationship identified here between CFR and target SpO2 may be more a relationship between CFR and national strategy. Target SpO2 may be more of an indicator of national policy.

### Limitations

This study highlights the variation in national guidelines for when to commence supplemental oxygen in patients with COVID-19. In of itself, this raises important questions as to the optimal response to COVID-19. Attempting to delineate the interventions and strategies that are potentially beneficial between nations is difficult without using a mortality estimation, which carries inherent confounders. CFR depends on many factors, not least of which is the accurate reporting of COVID-19 related deaths. Whilst we found no correlation between CFR and rates of testing or crude case burden, we could not account for disparities in reporting of deaths. We were aware of this limitation prior to the study, but agreed with the view that while CFR is unlikely to be numerically correct, the difference between countries will remain[3,4], and as such the association is likely to be accurate.

We undertook an analysis of the national guidelines using three independent investigators. The consensus amongst the investigators supports the accuracy of the target SpO2 extracted. The possibility remains that localities within a country, or individual doctors and nurses, chose not to follow their national guidelines. Even if such local differences were significant, the national guidelines *permits* not implementing oxygen therapy until the target oxygen level is reached, therefore triage systems, nurses and physicians can avoid admissions, and thus limit access to supplemental oxygen. Despite the prospect of local variations in following the guidelines, the presence of the guidelines will shape and likely reflect practice nationally.

## Conclusion

There is clear disparity between national guidelines for target oxygen saturations (SpO2) in COVID-19 across the countries analysed here, and such disparity is associated with national case fatality rates (CFR). Whilst there are multiple confounders to the CFR, the overall relationship between increasing CFR with a decreasing target SpO2 is likely real. The cause for the relationship may be a true causative effect of hypoxia on mortality. It may also be an indirect effect of delayed or reduced access to supplemental oxygen stemming from an overwhelmed or under-resourced health service, or, a similar policy-related reduction in access to supplemental oxygen associated with the differing overall national approaches to COVID-19 - stay home versus clinical assessment. All three possibilities highlighted here implicate delayed initiation of oxygen in the excess mortality associated with COVID-19.

Further research is needed to explore the population risk associated with delayed correction of hypoxia in COVID-19. Additionally, it would be useful to undertake a health-risk analysis, from a resource allocation perspective, on the benefits of increasing access to supplemental oxygen for patients with COVID-19 versus other interventions. As it stands currently, our results support the position that managing COVID-19 pneumonia should not differ from the management of other pneumonias, in so much as, access to supplemental oxygen is necessary to prevent excessive mortality.

## Data Availability

No additional data available for this study

## Contribution

DG, HD, AK, JN, SB and FB contributed to the conception and/or design of the study and contributed to the manuscript. DG, HD and FB conducted the analysis of the national guidelines. Statistical analysis was undertaken primarily by JN. DG wrote the majority of the manuscript. SB and FB undertook final edits. All contributors reviewed the final manuscript before submission.

## Supplementary File

**Supplementary Table 1.**
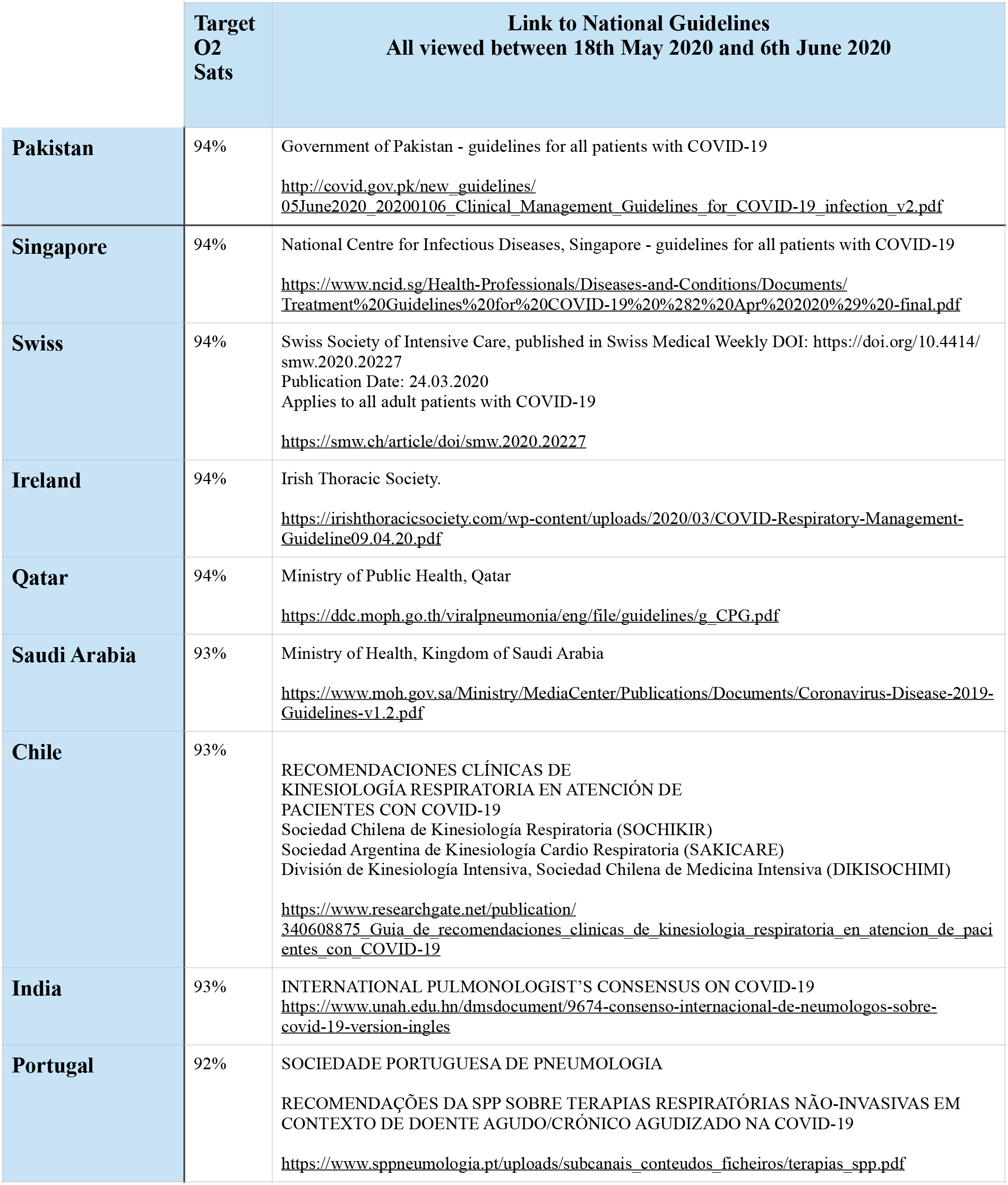
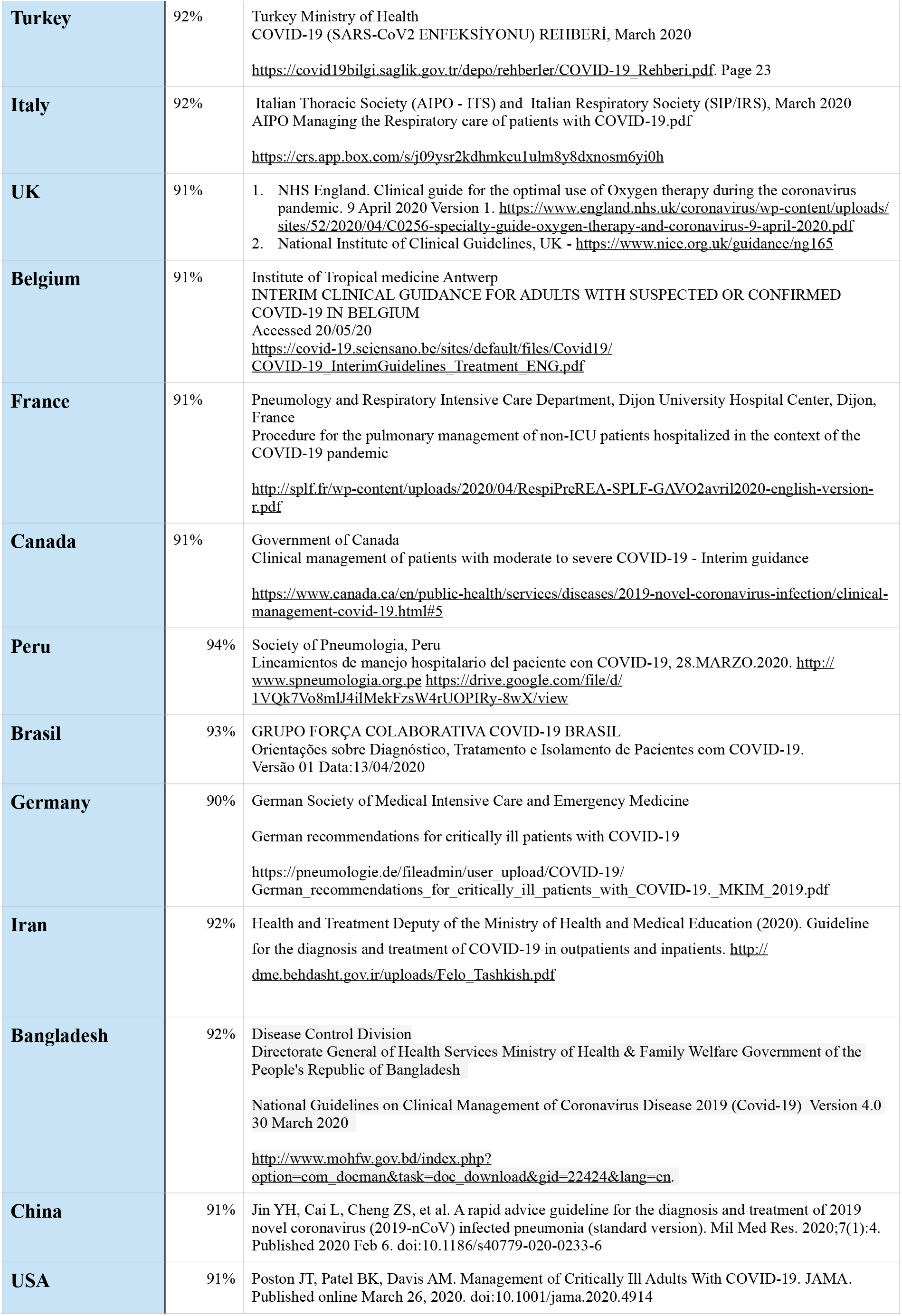
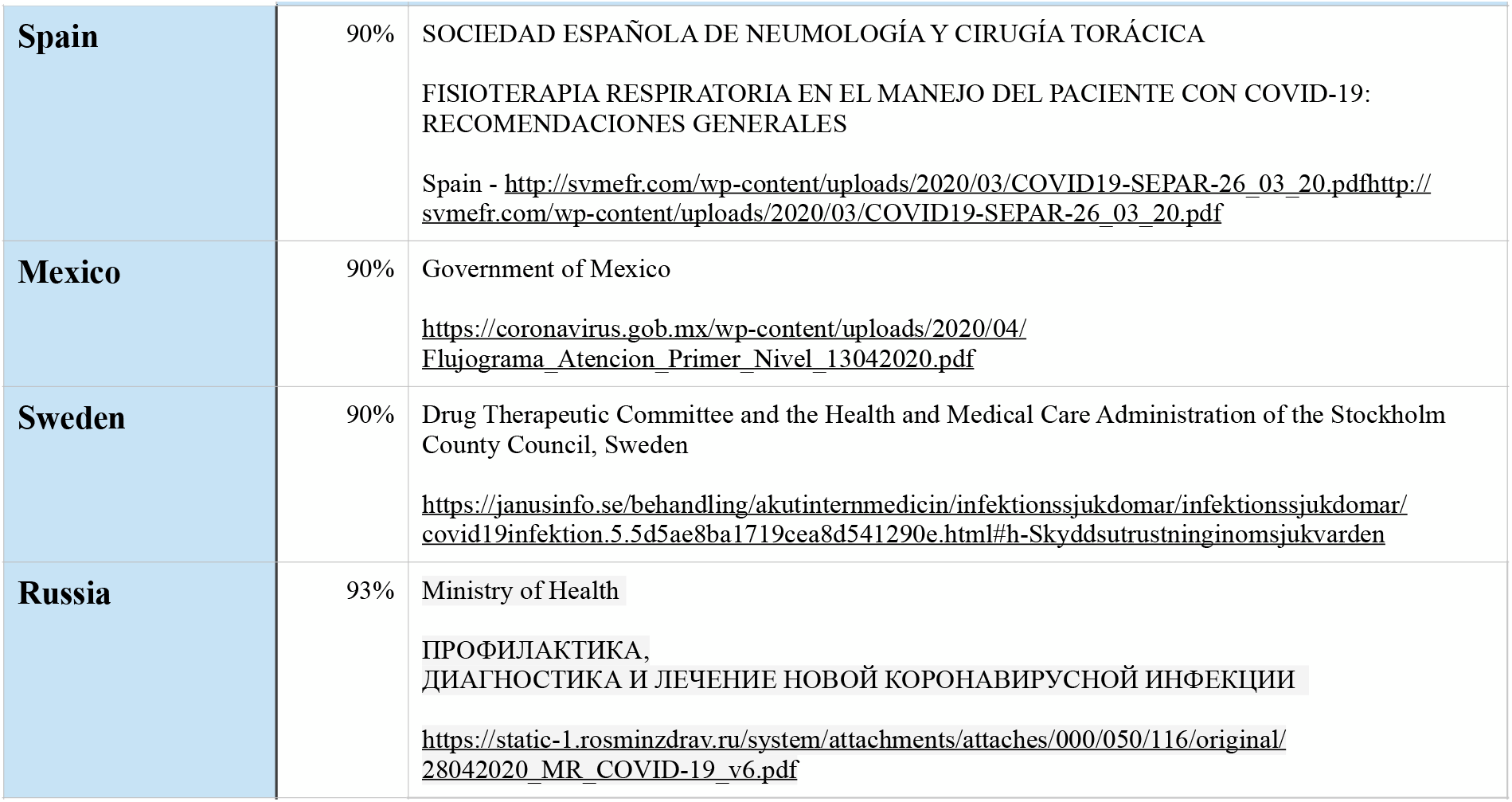
Target oxygen saturations with links to national guidelines.

